# Breast Cancer Risk Factors in the Abu Dhabi Population: A Retrospective Cohort Study

**DOI:** 10.1101/2025.02.13.25320817

**Authors:** Latifa Baynouna AlKetbi, Maha AlAzeezi, Rawan Ashoor, Nico Nagelkerke, Noura AlAlawi, Rudina AlKetbi, Hamda Aleissaee, Noura AlShamsi, Ahmed Humaid, Hanan Abdulbaqi, Toqa Fahmawee, Basil AlHashaikeh, Muna AlDobaee, Nayla AlAhbabi, AlYazia AlAzeezi, Fatima Shuaib, Jawaher Alnuaimi, Esraa Mahmoud, Mohammed AlMansoori, Sanaa AlKalbani, Wesayef AlDerie, Ekram Saeed, Amira AlAhmadi, Mohammad Sahyouni, Reem AlShamsi

## Abstract

**Background:** Breast cancer is the most common cancer in the United Arab Emirates (UAE), with the majority of cases characteristically occurring in women younger than 50 years. Studies on breast cancer risk factors in the UAE are few.

**Methods:** This retrospective study, from 2011 to 2013 until 2023, aimed to estimate the prevalence of a history of cancer and lifetime risk. A nested case-control study was designed from the cohort to study breast cancer risk with four controls for each case matched by the date of diagnosis.

**Results:** Breast cancer incidence was 0.85 %, 37 cases out of the 4338 females in the cohort. No males were diagnosed with the condition. Out of the 129 females with cancer, breast cancer was the most common type, 28% of all kinds of cancers.

Three risk factors for breast cancer were identified using conditional Cox regression analysis of the matched nested case-control study. The strongest was being a patient with diabetes Mellitus, HR=13.1 (3.2-52.8) P<0.001. Another metabolic abnormality that is associated with the occurrence of breast cancer in this population was a higher level of total cholesterol, HR=1.59 (1.03-2.44), P-value=0.035. Vitamin D showed a protective association with 58% reduced risk with levels above 75, HR=0.042 (0.002-0.73) P=0.042. The area under the ROC Curve for predicting breast cancer using this model was 0.753 (0.666-0.839). The area under the ROC Curve for predicting breast cancer using this model was 0.753 (0.666-0.839).

**Conclusion:** This study highlights the importance of understanding the various risk factors associated with breast cancer development and stresses the pivotal role of advancing diagnostic methods to improve early detection. By synthesizing current knowledge, this study aims to enhance our understanding of breast cancer’s complex nature and guide future research directions, screening practices, and preventive measurements.

## Introduction

Breast cancer was the most commonly diagnosed type of cancer globally in 2020. (Ref) It was the leading cause of cancer-related deaths among women and ranked as the fifth most common cause of cancer deaths overall (Ref). There is regional variation in incidence, with most regions in the world increasing since 1990, except North America. The Middle East and North Africa showed the most significant per-year increase in overall incidence. The global increase in BC incidence is seen in all age groups and is highest in women under 50 ^[1]^.. It is the most prevalent cancer in the United Arab Emirates (UAE), predominantly affecting women under 50 years of age, and it is the number one cause of cancer-related mortality in the UAE. One in five women with breast cancer in the UAE, 21.5%, are between the ages of 30 and 40 years ^[3]^.

With the increased incidence in the younger age groups, this variation in incidence is not, therefore, explained by increased life expectancy. Genetic predisposition through genetic mutations and environmental risk factors was found to play a key role. There are many genes found related to breast cancer, but other established risk factors are gender, age, estrogen hormone, family history, and unhealthy lifestyle.

The expression of these risk factors varies in different parts of the world. For example, the incidence of breast cancer is higher among younger females in Sudan, and advances in the early identification and prevention of breast cancer have gained great attention to improve survival and patients’ quality of life ^[2]^. Nevertheless, prevention of breast cancer extends to environmental as well as genetic factors. Therefore, studies in different geographical areas are of extreme importance and will impact various populations with different ethnicities and environmental characteristics.

The UAE has well-advanced healthcare services in managing breast cancer and follows the latest international guidelines and treatment modalities, including the availability of the latest drugs ^[3]^.

This retrospective cohort study is based on a national health screening program, and it focuses on breast cancer incidence, risk factors, and estimation of its risk in a population of the Arab population that was not well studied before.

## Methods

The cohort participants were 8699, who had their cardiovascular screening between 2011 and 2013 as part of the national program WEQAYA. This Abu Dhabi emirate-based program in the UAE targets UAE nationals over 18 to prevent and reduce the risks of cardiovascular diseases.

Among all participants, there were 64 who had cancer before the screening date; 37.5% were males and 62.5% were females. Cancer-free cases were 8635, 4338 of them were females. Follow-up of this population was done in 2023. Incidents of new cancer among the following population were collected. Data collected at the Outcome assessment was done in 2023; the average follow-up period was 9.2, with a minimum follow-up of less than a year and a maximum of 12 years. Data were collected by physicians and nurses who reviewed the Electronic Medical Records (EMR) of all subjects.

The study’s methodology was explained in other publications ()(). Variables collected at baseline included demographic data, self-reported health indicators including smoking status, physical activity, preexisting CVD, diabetes, hypertension, and dyslipidemia, and whether participants were taking medication for these conditions. Anthropometric measures included waist and hip circumference, body mass index (BMI in kg/m2), and a single arterial blood pressure reading. Hematological parameters included non-fasting glucose (mmol/L), total cholesterol, high-density lipoprotein (HDL) cholesterol (mmol/L), glycosylated hemoglobin (HbA1c), vitamin D, and creatinine (12). As there was missing data from some subjects, only subjects with completed data were included.

Breast cancer was further studied as it was the most prevalent, with 37 breast cancer cases. The design was a nested case-control study. Cases were matched with four controls for each case based on the year the breast cancer was diagnosed. Controls for each case had the same follow-up period. This study design of a nested matched case-control study was used to study possible determinants of the disease.

### Statistical analysis

Frequencies, descriptive statistics, and Cox survival regression with all types of cancer and breast cancer events as endpoints.

## Results

Among participants in the national screening program in 2011-2013 there were 129 out of the 4338 female participants had cancer, which represents 2.9%. Breast cancer is the highest among female cancers and has a prevalence that exceeds all types of cancer in both genders, Table 2. With regards to breast cancer, it shows that the average for new incident cases was 43.9 years. Breast cancer cases had more prevalence of diabetes at baseline and more hypertension prevalence. They had higher average cholesterol levels and BMI, 4.8 compared to 5.1 and 28.9 compared to 30.3, respectively.

Three risk factors for breast cancer were identified using conditional Cox regression analysis of the matched nested case-control study. The strongest was being a patient with diabetes Mellitus, HR=13.1 (3.2-52.8) P<0.001. Another metabolic abnormality that is associated with the occurrence of breast cancer in this population was a higher level of total cholesterol, HR=1.59 (1.03-2.44), P-value=0.035. Vitamin D showed a protective association with 58% reduced risk with levels above 75, HR=0.042 (0.002-0.73) P=0.042. with univariable conditional Cox regression, table 2-b, only Diabetes had a significant association. Age was not significant with the univariable conditional Cox regression as well. Figure 1 The Kapplan-Meir analysis shows the influence of vitamin D status and diabetes mellitus on the occurrence of breast cancer over the follow-up period, Figures 2 A and B, respectively.

**Figure 1.**
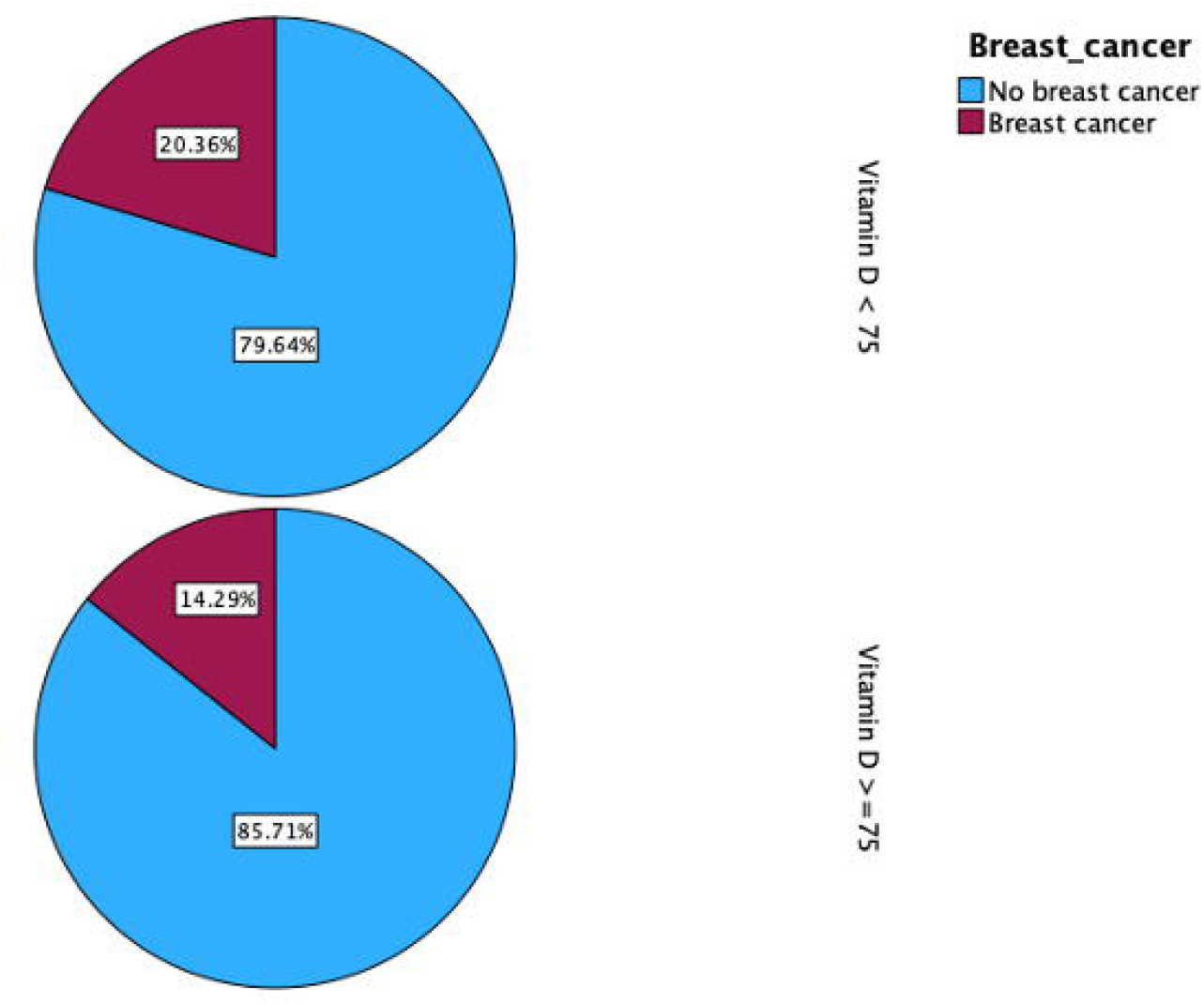
Incidence of breast cancer among women participating in the study distributed by vitamin D status at the start of the study.

**Figure 2.**
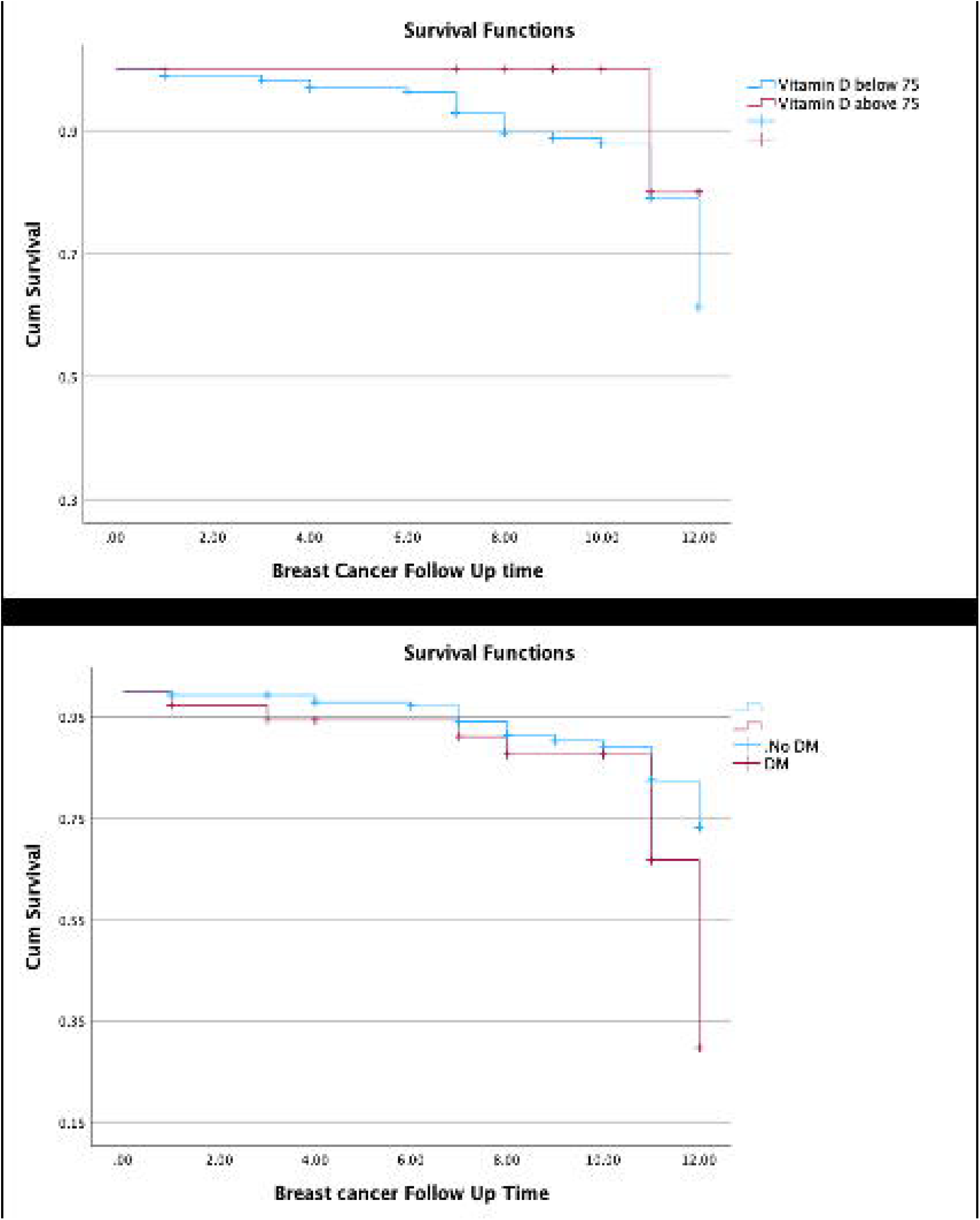
Kapplan-Meir analysis of the incidence of breast cancer in relation to vitamin D status Figures 2 A and diabetes mellitus Figures 2 B over the follow-up period.

The area Under the ROC Curve for predicting breast cancer using this model was 0.703 (0.612-0.793). Figure 3

**Figure 3.**
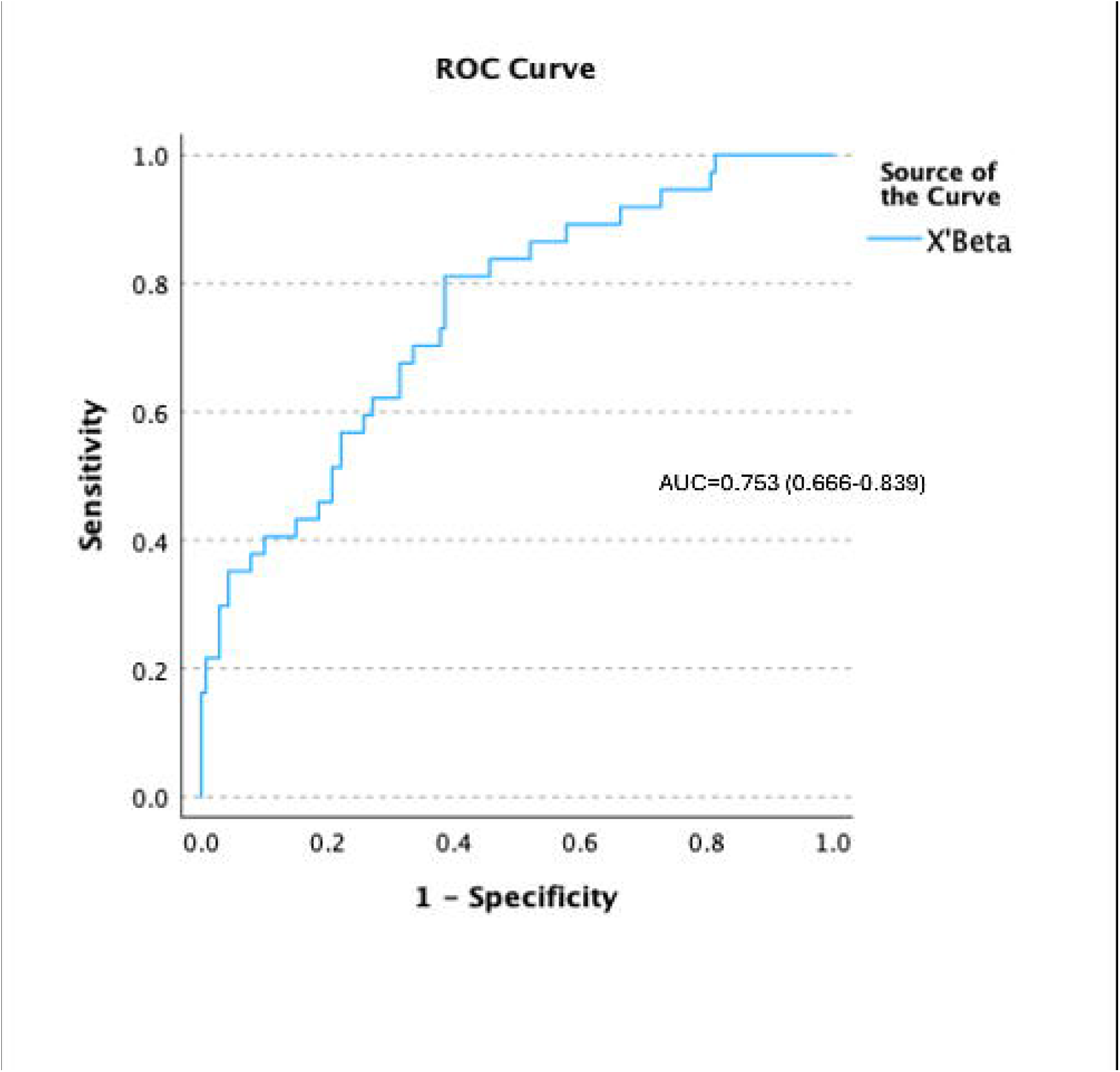
The area Under the ROC Curve for predicting breast cancer using the developed model.

## Discussion

Age was not a significant risk factor in increasing the risk of breast cancer among this sample of the population, although it is a well-established risk factor [4]. No other study has shown a similar lack of age influence on the risk of breast cancer. A possible explanation is that while each generation experiences an increase with age, this is offset by a generation effect, with the younger generation having a higher risk than older generations due to changes in lifestyles and other social factors. For example, earlier, more traditional generations had higher fertility and practiced longer breastfeeding periods ^**[5]**^. In addition, diet changes and less physical activity may have resulted in metabolic abnormalities, such as diabetes, high cholesterol, and low vitamin D, at younger ages, which increased breast cancer risk. Nevertheless, other known risk factors, such as obesity, were not significant determinants, perhaps due to a lack of statistical power.

High cholesterol has emerged as a strong predictor of the risk of breast cancer. Lipids’ role in cancer epidemiology is receiving increasing attention, and multiple mechanisms for cholesterol-mediated cancer progression have been proposed. Notably, cancer cells are dependent on cholesterol. Malignant progression is associated with high cellular demand for cholesterol, and extracellular cholesterol uptake is often elevated in cancer cells to meet their metabolic needs. It has been suggested that ^[6]^ manipulation of their levels could potentially provide novel, efficient therapeutic strategies for treatment ^[7]^. The SCARB1 gene also encodes the scavenger receptor class B type I (SR-BI), which is important in regulating cholesterol exchange between cells and high-density lipoproteins. Solid tumors have been shown to accumulate more cholesterol than the host healthy tissues. Although no study examining the role of SR-BI in the promotion of cell transformation has been performed, it has been recognized that cancer cells can use the HDL/SR-BI pathway to take up cholesteryl ester and enhance malignant phenotypes ^[8] [9]^.

The role of diabetes as a risk factor for breast cancer is worrisome. While this is a known risk factor, our estimates of its relative risk are particularly high. Also, the UAE is among the countries with the highest prevalence of DM2, which may have major public health implications ^[10]^. Related risk factors explored include hyperinsulinemia, elevated IGFs, hyperglycemia, dyslipidemia, adipokines, inflammatory cytokines, and the gut microbiome ^[11]^. Modification of such risk factors, e.g., through intentional weight loss, may protect against cancer development, and therapies for diabetes may prove to be effective adjuvant agents in reducing cancer incidence and progression ^[12] [13]^.

Additionally, patients with diabetes were found to have higher mortality from breast cancer due to the Warburg effect ^[14]^. Interestingly, an oncogene, the P4HA3 gene, which is significantly upregulated in breast cancer, was also significantly upregulated in the subcutaneous adipose tissue of obese and type 2 diabetes mellitus (T2DM) patients, with a functional role in the differentiation of adipocytes and insulin resistance. The high prevalence of diabetes and obesity in the UAE may thus increase the cancer risk in this population ^[15] (16)^.

Another risk factor in this cohort is vitamin D deficiency, confirming an earlier. Systematic review and meta-analysis, in which 25(OH)D deficiency was directly related to breast cancer ^[17]^. This was further supported by an Umbrella Review on the Relationship between Vitamin D Levels and Cancer ^[18]^. This review showed inverse correlations between 25(OH)D levels and mortality and mostly inverse correlations between 25(OH)D levels and incidence ^[18]^. Hypothesized that VDR gene variation shapes the human microbiome and that VDR deficiency leads to dysbiosis. Intestinal VDR dysfunction and gut dysbiosis lead to a high risk of extraintestinal tumorigenesis ^[19]^. Correlating microbiome and carcinogenesis is important as it can provide insights into the mechanisms by which microbial dysbiosis can influence cancer development and progression, suggesting the use of the microbiome as a potential tool for prognostication and personalized therapy ^[20]^.

The etiology of breast cancer and cancer, in general, is complex and involves both additional lifestyle ^[21] [22] (23)^ and genetic factors. An ongoing genomic study in the UAE is expected to knowledge in this field ^[24]^.

The limitations of this study include a relatively small sample size and the absence of data on other significant risk factors, such as positive family history, reproductive history, and hormone replacement history. However, a major strength of the study is that it is a large community-based cohort with a long follow-up period.

## Conclusion

Increased HDL levels and older age were the identified risk factors when all cancer types were studied together. Diabetes and hypercholesterolemia were identified risk factors for breast cancer in this cohort, while vitamin D levels of 75 or more are protective from developing breast cancer. This study calls for future efforts that should prioritize expanding national registries and collecting clinically relevant variables. This can enhance understanding of the clinical and molecular profile of breast cancer in the country and, crucially, provide survival metrics that can guide management strategies.

## Data Availability

All data produced in the present study are available upon reasonable request to the authors

## Declarations

### Ethical approval and consent to participate

The study was approved by the AlAin Human Ethics Committee, approval number 13/58, and Ambulatory Healthcare Services IRB 19-2022. All methods were carried out under relevant guidelines and regulations. The authors confirm that the study was conducted in accordance with the Helsinki Declaration.

### Consent statement in the Ethics approval and consent to participate

Informed consent was waived by the IRBs as the study was designed for retrospective data gathered as part of patient care and anonymized at analysis.

### Authors’ contributions

LBK and NN conceptualized and analyzed data. LBK, MAA, and RA wrote the manuscript; all other co-authors collected data and reviewed the manuscript. All authors have read and approved the final manuscript.

## Acknowledgments

None.

## Consent to publish

Not Applicable.

### Availability of data and materials

Data availability is restricted due to institution policies.

## Reference

Global breast cancer incidence and mortality trends by region, age-groups, and fertility patterns—eClinicalMedicine. (n.d.). Retrieved November 18, 2024, from https://www.thelancet.com/journals/eclinm/article/PIIS2589-5370(21)00265-0/fulltext

Sun, Y.-S., Zhao, Z., Yang, Z.-N., Xu, F., Lu, H.-J., Zhu, Z.-Y., Shi, W., Jiang, J., Yao, P.-P., & Zhu, H.-P. (2017). Risk Factors and Preventions of Breast Cancer. International Journal of Biological Sciences, 13(11), 1387–1397. 10.7150/ijbs.21635

Breast Cancer in the United Arab Emirates | JCO Global Oncology. (n.d.-a). Retrieved November 18, 2024, from https://ascopubs.org/doi/10.1200/GO.22.00247

Ł ukasiewicz, S., Czeczelewski, M., Forma, A., Baj, J., Sitarz, R., & Stanisławek, A. (2021). Breast Cancer—Epidemiology, Risk Factors, Classification, Prognostic Markers, and Current Treatment Strategies—An Updated Review. Cancers, 13(17), 17. https://doi.org/10.3390/cancers13174287

Patterns of breastfeeding practices among infants and young children in Abu Dhabi, United Arab Emirates | International Breastfeeding Journal | Full Text. (n.d.). Retrieved November 18, 2024, from https://internationalbreastfeedingjournal.biomedcentral.com/articles/10.1186/s13006-018-0192-7

Why make it if you can take it: Review on extracellular cholesterol uptake and its importance in breast and ovarian cancers | Journal of Experimental & Clinical Cancer Research | Full Text. (n.d.). Retrieved November 18, 2024, from https://jeccr.biomedcentral.com/articles/10.1186/s13046-024-03172-y

Frontiers | SR-BI: Linking Cholesterol and Lipoprotein Metabolism with Breast and Prostate Cancer. (n.d.). Retrieved November 18, 2024, from https://www.frontiersin.org/journals/pharmacology/articles/10.3389/fphar.2016.00338/full

Nam, S. Y., Jo, J., & Cho, C.-M. (2024). A population-based cohort study of longitudinal change of high-density lipoprotein cholesterol impact on gastrointestinal cancer risk. Nature Communications, 15(1), 2923. 10.1038/s41467-024-47193-9

Deficiency of SR□ B1 reduced the tumor load of colitis□ induced or APCmin/+□ induced colorectal cancer—Chen—2023—Cancer Medicine—Wiley Online Library. (n.d.). Retrieved November 18, 2024, from https://onlinelibrary.wiley.com/doi/10.1002/cam4.6534

Type 2 Diabetes and Subsequent Incidence of Breast Cancer in the Nurses’ Health Study | Diabetes Care | American Diabetes Association. (n.d.). Retrieved November 18, 2024, from https://diabetesjournals.org/care/article/26/6/1752/26368/Type-2-Diabetes-and-Subsequent-Incidence-of-Breast

Diabetes, Obesity, and Breast Cancer | Endocrinology | Oxford Academic. (n.d.). Retrieved November 18, 2024, from https://academic.oup.com/endo/article/159/11/3801/5094963

Obesity and Diabetes: The Increased Risk of Cancer and Cancer-Related Mortality | Physiological Reviews. (n.d.). Retrieved November 18, 2024, from https://journals.physiology.org/doi/full/10.1152/physrev.00030.2014

Biological Basis of Breast Cancer-Related Disparities in Precision Oncology Era. (n.d.). Retrieved November 18, 2024, from https://www.mdpi.com/1422-0067/25/7/4113

Eketunde AO. Diabetes as a Risk Factor for Breast Cancer. Cureus. 2020 May 7;12(5):e8010. doi: 10.7759/cureus.8010. PMID: 32528752; PMCID: PMC7279688.

United Arab Emirates diabetes report 2000—2045. (n.d.-b). Retrieved November 18, 2024, from https://diabetesatlas.org/data/

Neagu, A.-N., Bruno, P., Johnson, K. R., Ballestas, G., & Darie, C. C. (2024). Biological Basis of Breast Cancer-Related Disparities in Precision Oncology Era. International Journal of Molecular Sciences, 25(7), Article 7. 10.3390/ijms25074113

Hossain, S., Beydoun, M. A., Beydoun, H. A., Chen, X., Zonderman, A. B., & Wood, R. J. (2019). Vitamin D and breast cancer: A systematic review and meta-analysis of observational studies. Clinical Nutrition ESPEN, 30, 170–184. 10.1016/j.clnesp.2018.12.085

Umbrella Review on the Relationship between Vitamin D Levels and Cancer. (n.d.). Retrieved November 18, 2024, from https://www.mdpi.com/2072-6643/16/16/2720

Zhang, Y.-G., Xia, Y., Zhang, J., Deb, S., Garrett, S., & Sun, J. (2023). Intestinal vitamin D receptor protects against extraintestinal breast cancer tumorigenesis. Gut Microbes, 15(1), 2202593. 10.1080/19490976.2023.2202593

Jotshi, A., Sukla, K. K., Haque, M. M., Bose, C., Varma, B., Koppiker, C. B., Joshi, S., & Mishra, R. (2023). Exploring the human microbiome – A step forward for precision medicine in breast cancer. Cancer Reports, 6(11), e1877. https://doi.org/10.1002/cnr2.1877

Physical Exercise and the Hallmarks of Breast Cancer: A Narrative Review. (n.d.). Retrieved November 18, 2024, from https://www.mdpi.com/2072-6694/15/1/324

Association between sleep traits and risk of colorectal cancer: A bidirectional Mendelian randomization study—He—Journal of Gastrointestinal Oncology. (n.d.). Retrieved November 18, 2024, from https://jgo.amegroups.org/article/view/88579/html

Brown, R. B., Bigelow, P., & Dubin, J. A. (2023). Breast Cancer and Bone Mineral Density in a U.S. Cohort of Middle-Aged Women: Associations with Phosphate Toxicity. Cancers, 15(20), Article 20. 10.3390/cancers15205093

The Emirati Genome Programme | The Official Portal of the UAE Government. (n.d.). Retrieved November 18, 2024, from https://u.ae/en/information-and-services/health-and-fitness/research-in-the-field-of-health/the-emirati-genome-programme

