## Supplementary material for "Breast Cancer Risk Factors in the Abu Dhabi Population: A Retrospective Cohort Study": Table 1: Table 1 Breast cancer .docx

Table 1 Breast cancer and non-breast cancer subjects’ characteristics

| Breast cancer | Did not develop breast cancer | | Breast cancer | |
| --- | --- | --- | --- | --- |
|  | Mean | SD | Mean | SD |
| Whole_age | 39.0 | 14.7 | 43.9 | 12.0 |
| SBP | 116.0 | 15.3 | 117.7 | 17.2 |
| DBP | 70.6 | 9.3 | 72.1 | 10.5 |
| cholesterol_value | 4.8 | 1.1 | 5.1 | 1.6 |
| HBA1C | 5.7 | 0.7 | 5.9 | 1.0 |
| hdl | 1.4 | 0.4 | 1.4 | 0.4 |
| VITAMIN_D | 36.1 | 23.1 | 40.2 | 20.3 |
| bmi1 | 28.9 | 6.7 | 30.3 | 6.5 |
| GFR | 112.7 | 18.7 | 106.0 | 19.3 |
|  | No. | % | No. | % |
| X_Smok_Screen | 0 | 0 | 0 | 0 |
| CurrentSmoker | 1 | 0.7 | 1 | 2.7 |
| DM_BEFORE_SCRN | 23 | 15.5 | 14 | 37.8 |
| HTN_BEFORE_SCRN | 24 | 16.2 | 9 | 24.3 |
| CHD_BEFORE | 3 | 2 | 1 | 2.7 |

| Ocuarance of breast cancer in the cohort | | | | |
| --- | --- | --- | --- | --- |
| With breast cancer After | 41 | 0.9 | 1 | 0 |
| With breast cancer Befor | 11 | 0.3 | 0 | 0 |
