## Supplementary material for "Breast Cancer Risk Factors in the Abu Dhabi Population: A Retrospective Cohort Study": Table 2: Table 2 .docx

Table 2 Risk factors of breast cancer.

A. Multivariable conditional Cox regression

|  | B | P value | HR | 95.0% CI for HR | |
| --- | --- | --- | --- | --- | --- |
| DM before screening | 2.572 | <.001 | 13.088 | 3.243 | 52.816 |
| Total Cholesterol | 0.463 | 0.035 | 1.588 | 1.033 | 2.443 |
| Age | -0.002 | 0.959 | 0.998 | 0.908 | 1.096 |
| Vitamin D | 0.028 | 0.411 | 1.029 | 0.962 | 1.1 |
| Vitamin D >=75 | -3.182 | 0.03 | 0.042 | 0.002 | 0.736 |
| Height | -0.053 | 0.177 | 0.948 | 0.878 | 1.024 |
| SBP | 0 | 0.994 | 1 | 0.968 | 1.033 |
| FAM_CANCER_HIS | -0.704 | 0.456 | 0.495 | 0.078 | 3.154 |
| bmi1 | -0.021 | 0.567 | 0.98 | 0.913 | 1.051 |
| Cancer before (not Breast) | 11.743 | 0.969 | 125806.863 | 0 | 1.25E+260 |
| HTN_BEFORE_SCRN | -0.442 | 0.518 | 0.643 | 0.168 | 2.456 |
| WHR | -3.955 | 0.18 | 0.019 | 0 | 6.195 |
| WALKING_STAT | -0.612 | 0.282 | 0.542 | 0.178 | 1.655 |
| GFR | -0.017 | 0.448 | 0.983 | 0.942 | 1.027 |
| housewife | 0.649 | 0.272 | 1.913 | 0.602 | 6.085 |
| EDUCATION_LVL | 0.216 | 0.305 | 1.241 | 0.821 | 1.876 |
| VITAMIN_D*Age | 0 | 0.951 | 1 | 0.998 | 1.002 |

B. Univariable conditional Cox regression

|  | | B | P value | HR | 95.0% CI for HR | |
| --- | --- | --- | --- | --- | --- | --- |
| DM | 1.261 | | 0.004 | 3.531 | 1.513 | 8.237 |
| HTN | 0.499 | | 0.256 | 1.647 | 0.696 | 3.901 |
| Vitamin more than 75 | -0.515 | | 0.508 | 0.598 | 0.13 | 2.747 |
| Vitamin D | 0.008 | | 0.32 | 1.008 | 0.993 | 1.023 |
| cholesterol_value | 0.181 | | 0.172 | 1.198 | 0.924 | 1.554 |
| hdl | 0.252 | | 0.609 | 1.287 | 0.489 | 3.387 |
